# Demographic and clinical characteristics of patients with functional motor disorders: the prospective Salpêtrière cohort

**DOI:** 10.1101/2021.02.04.21251123

**Authors:** Béatrice Garcin, Nicolas Villain, Francine Mesrati, Lionel Naccache, Emmanuel Roze, Bertrand Degos

## Abstract

Functional motor disorders (FMD) are common and disabling. They are known to affect predominantly women and to start at young or middle age but to date, large case series are lacking, and demographic and clinical characteristics of patients with FMD rely on data from small cohorts. The current study aimed at describing the demographic and clinical characteristics of a large cohort of FMD patients.

**Methods:** We prospectively collected data from FMD patients who were referred to the Neurophysiology Department of the Pitié-Salpêtrière University Hospital between 2008 and 2016 for treatment with repeated transcranial magnetic stimulation.

**Results:** 482 patients were included. There was a majority of women (73.7%) with a median age of 40 years old at TMS treatment. Median age at symptoms onset was 35.5 years old and symptoms were mostly characterized by an acute (47.3%) or subacute (46%) onset. Only 23% of patients were active workers while 58.3% were unemployed for medical reasons. Half of the patients suffered from functional motor weakness (n= 241) and the other half suffered from movement disorders (n=241), mainly represented by tremor (21.15%) and dystonia (20.5%). 33.6% had no psychiatric comorbidity and 17.4% reported no history of trauma. No significant differences were found in age or gender according to clinical phenotypes.

**Conclusion:** We present the largest cohort of patients with FMD to date. This cohort will contribute to a better understanding of FMDs and their risk factors.

## Introduction

Functional Neurological Disorders (FND)s are the second commonest reason for referral in neurology outpatient clinics (Stone et al. 2009). FND can affect motor and/or sensory functions, other senses, cognition, and include non-epileptic seizures. The current understanding of the disease involves brain network dysfunction that is potentially reversible, individual vulnerability and the possibility of an emotional/psychological trigger. Functional motor disorder (FMD) is a common presentation of FND. Patients present with a variety of symptoms of altered movement, including weakness, tremor, jerks, stereotypies, dystonia and parkinsonism, often displaying combinations of multiple phenotypes. FMD is known to predominantly affect women and to start at young to middle age (Baizabal-Carvallo et Jankovic 2020; Tinazzi et al. 2020; Binzer, Andersen, et Kullgren 1997; Schrag et al. 2004; Factor, Podskalny, et Molho 1995; Jon Stone, Warlow, et Sharpe 2010). Recent studies have suggested that some phenotypes affect more one gender or one age range. For example, previous studies suggested that functional myoclonus affects more men (55%)(van der Salm et al. 2014), while functional parkinsonism is more frequent in older patients (Ambar Akkaoui et al. 2020). Some controversies also exist about FMD affecting more the left side of the body (J. Stone et al. 2002). However, large case series are lacking and most studies are small case series of specific phenotypes. They are necessary to better characterize the specificities of this patient population, and to draw hypotheses about the underlying risk factors for developing FMD. Additionally, this patient population is young or middle-aged, and there is a lack of data about work status and the economic burden of the disease.

In the present study, we aimed at describing the demographic and clinical characteristics of a prospective patient population suffering from FMD, and at assessing the repercussions of FMD on work status. Our secondary aim was to assess whether patients’ population differ within distinct clinical phenotypes.

## Material & Methods

### Population

We collected demographic and clinical data of all consecutive patients addressed to the Neurophysiology Department of Pitié-Salpêtrière University Hospital for functional motor disorders (FMD) over a period of 8 years. All patients were referred for transcranial magnetic stimulation (TMS) as a treatment of FMD (Garcin et al. 2013). A thorough clinical investigation in the Movement Disorder Unit of the Pitié-Salpêtrière University Hospital was performed for all patients. The diagnosis of functional motor weakness was performed by a neurologist from the Neurology Department of Pitié-Salpêtrière University Hospital after exhaustive investigations, following DSM5 criteria (« APA - Diagnostic and Statistical Manual of Mental Disorders DSM-5 Fifth Edition » s. d.). Functional movement disorders were diagnosed by a movement disorders expert (BD or ER) following the criteria for clinically established functional movement disorders, as defined by Gupta and Lang (Gupta et Lang 2009). The data collection was approved by the National Commission on Informatics and Liberty (CNIL) and was approved by the local ethics committee (CPP IdF 6, Pitié-Salpêtrière University Hospital).

### Data collection & analysis

Clinical and demographic data were collected from patients’ medical records and completed by a systematic medical interview before TMS treatment (FM). Age at symptoms onset, age at TMS treatment, gender, level of education, medical history and psychiatric comorbidities were collected. The onset of symptoms was defined as follows: sudden if the patient reported a sudden onset of symptoms at a precise moment, subacute if symptoms had settled progressively across several weeks to 3 months, chronic if symptoms appeared progressively upon more than 3 months. History of trauma was either spontaneously reported by the patient or reported during the medical interview. A psychotrauma was defined as a painful memory of an unexpected event that was reported to be traumatic by the patient (e.g. psychological harassment, death of a relative, diagnosis announcement…). A physical trauma was defined as a painful memory of a physical integrity violation reported as being traumatic by the patient (e.g. accident, surgery…), with or without physical sequelae. The occupational status was defined as active if the patient was working or active in another way (e.g.: studies); the absence of activity was then either classified as being unemployed due to non-medical reasons (e.g. unemployment, retirement…) or as being unemployed due to medical reasons (injury-on-duty leave, sick leave, disability living allowance).

Movement disorder phenomenology was classified for each patient by a movement disorder specialist (BD and ER). In the case of multiple functional symptoms, the main symptom was used for classification and the topography of symptoms was also reported.

Statistical analyses were conducted using SPSS software version 23 (http://www-01.ibm.com/software/analytics/spss/), with a threshold set at p<0.05 for significance. Data were first reported for the whole group of patients. Then, we compared the characteristics of patients with the following motor phenotypes: functional motor weakness versus movement disorders and then across different movement disorders subtypes using Student t-test, Chi^2^ test and ANOVAs.

## Results

### Whole patients group

Over a period of 8 years, 485 patients were referred to the Neurophysiology Department of the Pitié-Salpêtrière University Hospital for TMS treatment of FMD. Three patients were excluded because the motor and sensory evoked potentials were abnormal and induced diagnosis reconsideration; 482 patients were included in the present study. Demographic and clinical characteristics of patients are described in Table 1 and figure 1 and 2. There was a majority of women (73.7%) with a median age of 40 years old (range 8-77) at TMS treatment. Median age at the symptoms onset was 35.5 years old (range 3-75) and symptoms were mostly characterized by an acute (47.3%) or subacute (46%) onset. Only 23% of patients were active workers while 58.3% were unemployed for medical reasons. Regarding the phenomenology of symptoms, half of the patients referred for TMS treatment suffered from functional motor weakness (n= 241) and the other half suffered from movement disorders (n=241), mainly represented by tremor (21.15%) and dystonia (20.5%). The other movement disorders were jerky dystonia (4,6%), stereotypies (1,9%) and parkinsonism (1,9%). Amongst our patients, 18.8% had a medical history of other neurological disorders, 24.7% of orthopaedic-rheumatological disorders, complex regional pain syndrome or fibromyalgia, 13.3% had a history of abdominal or pelvic disorder. 47.3% of patients had no somatic medical history. Regarding psychiatric comorbidities, 52.7% of patients suffered from mood disorders and 33.6% had no psychiatric comorbidity. Finally, 50.6% of the patients reported a history of psychological trauma, 20.1% a physical trauma without sequelae and 8.7% surgical procedures, while 17.4% reported no history of trauma.

**Table 1.**
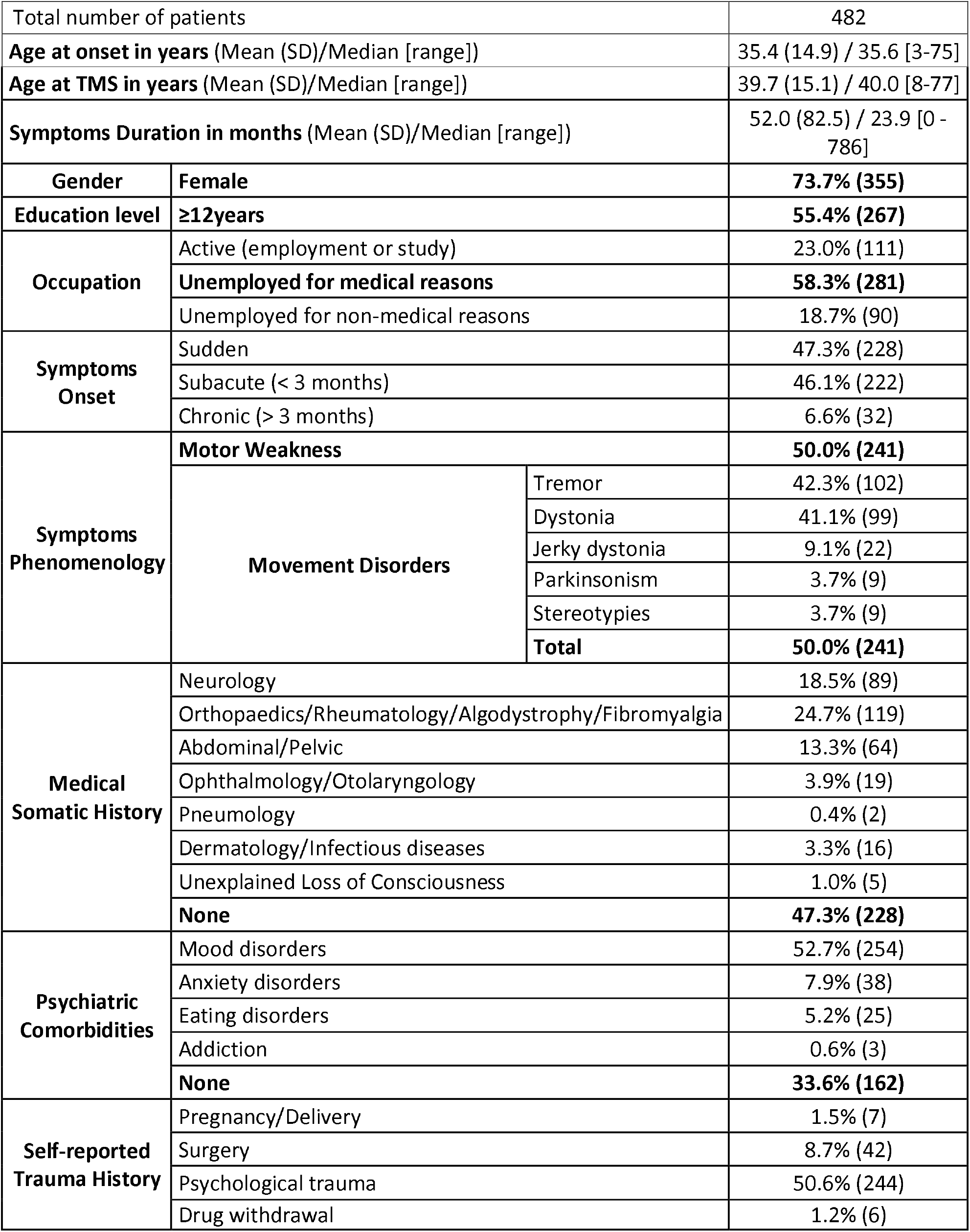
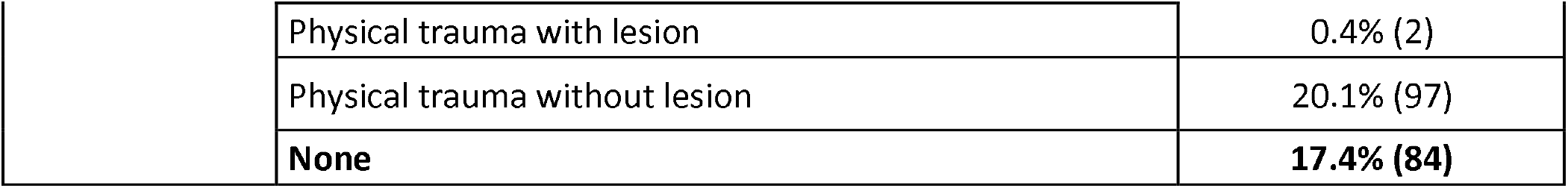
Demographical, clinical characteristics and medical history of patients with functional motor disorders.

|  |  |  |  |
| --- | --- | --- | --- |
| Total number of patients |  |  | 482 |
| Age at onset in years (Mean (SD))/Median [range]] |  |  | 35.4 (14.9) / 35.6 [3-75] |
| Age at TMS in years (Mean (SD))/Median [range]] |  |  | 39.7 (15.1) / 40.0 [8-77] |
| Symptoms Duration in months (Mean (SD))/Median [range]] |  |  | 52.0 (82.5) / 23.9 [0 - 786] |
| Gender | Female |  | 73.7% (355) |
| Education level | ≥12years |  | 55.4% (267) |
| Occupation | Active (employment or study) |  | 23.0% (111) |
|  | Unemployed for medical reasons |  | 58.3% (281) |
|  | Unemployed for non-medical reasons |  | 18.7% (90) |
| Symptoms Onset | Sudden |  | 47.3% (228) |
|  | Subacute (< 3 months) |  | 46.1% (222) |
|  | Chronic (> 3 months) |  | 6.6% (32) |
| Symptoms Phenomenology | Motor Weakness |  | 50.0% (241) |
|  | Movement Disorders | Tremor | 42.3% (102) |
|  |  | Dystonia | 41.1% (99) |
|  |  | Jerky dystonia | 9.1% (22) |
|  |  | Parkinsonism | 3.7% (9) |
|  |  | Stereotypies | 3.7% (9) |
|  |  | Total | 50.0% (241) |
| Medical Somatic History | Neurology |  | 18.5% (89) |
|  | Orthopaedics/Rheumatology/Algodystrophy/Fibromyalgia |  | 24.7% (119) |
|  | Abdominal/Pelvic |  | 13.3% (64) |
|  | Ophthalmology/Otolaryngology |  | 3.9% (19) |
|  | Pneumology |  | 0.4% (2) |
|  | Dermatology/Infectious diseases |  | 3.3% (16) |
|  | Unexplained Loss of Consciousness |  | 1.0% (5) |
|  | None |  | 47.3% (228) |
| Psychiatric Comorbidities | Mood disorders |  | 52.7% (254) |
|  | Anxiety disorders |  | 7.9% (38) |
|  | Eating disorders |  | 5.2% (25) |
|  | Addiction |  | 0.6% (3) |
|  | None |  | 33.6% (162) |
| Self-reported Trauma History | Pregnancy/Delivery |  | 1.5% (7) |
|  | Surgery |  | 8.7% (42) |
|  | Psychological trauma |  | 50.6% (244) |
|  | Drug withdrawal |  | 1.2% (6) |

|  |  |  |
| --- | --- | --- |
|  | Physical trauma with lesion | 0.4% (2) |
|  | Physical trauma without lesion | 20.1% (97) |
|  | **None** | **17.4% (84)** |

**Figure 1.**
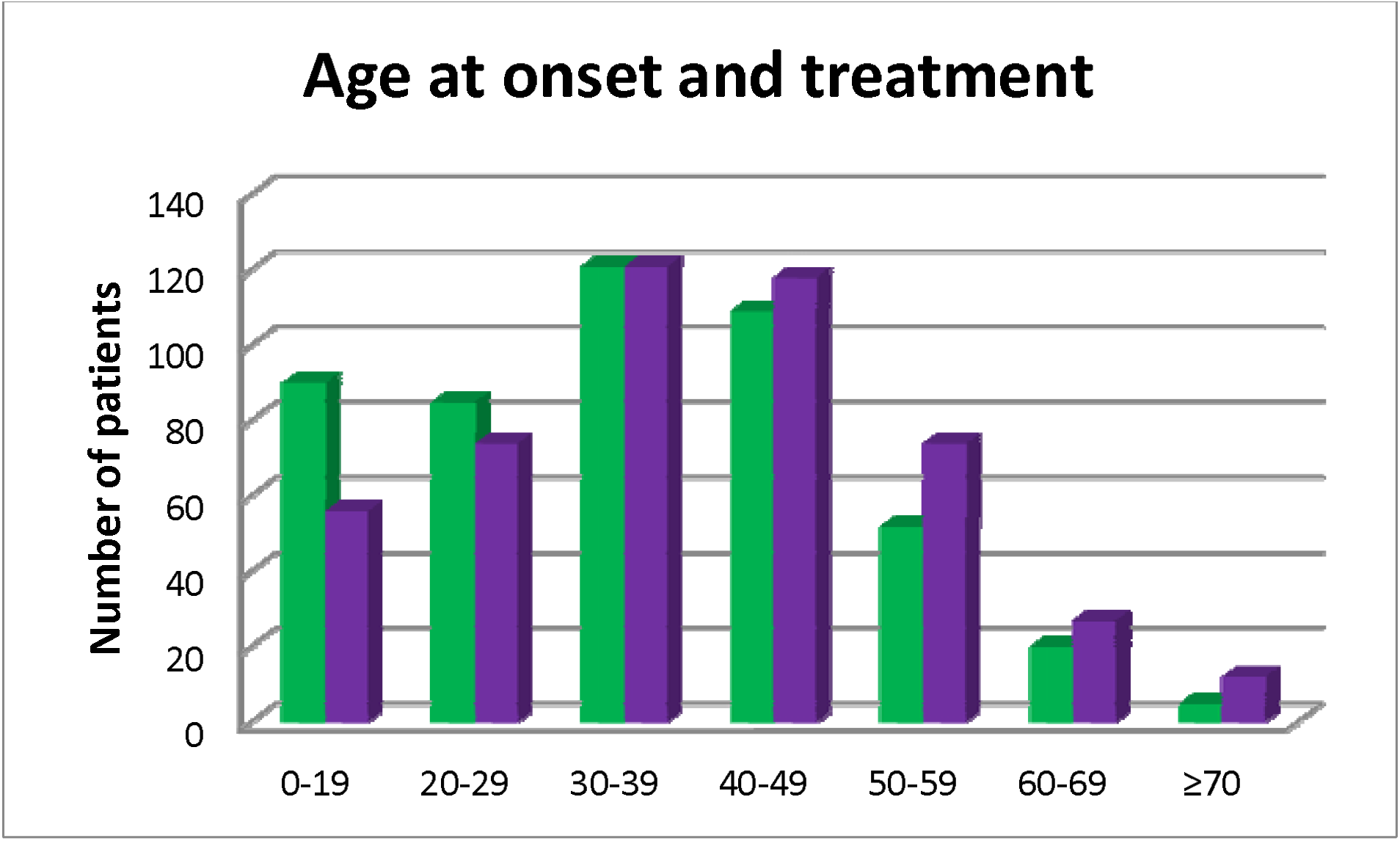
Age at symptom onset in green and at TMS treatment in purple.

**Figure 2.**
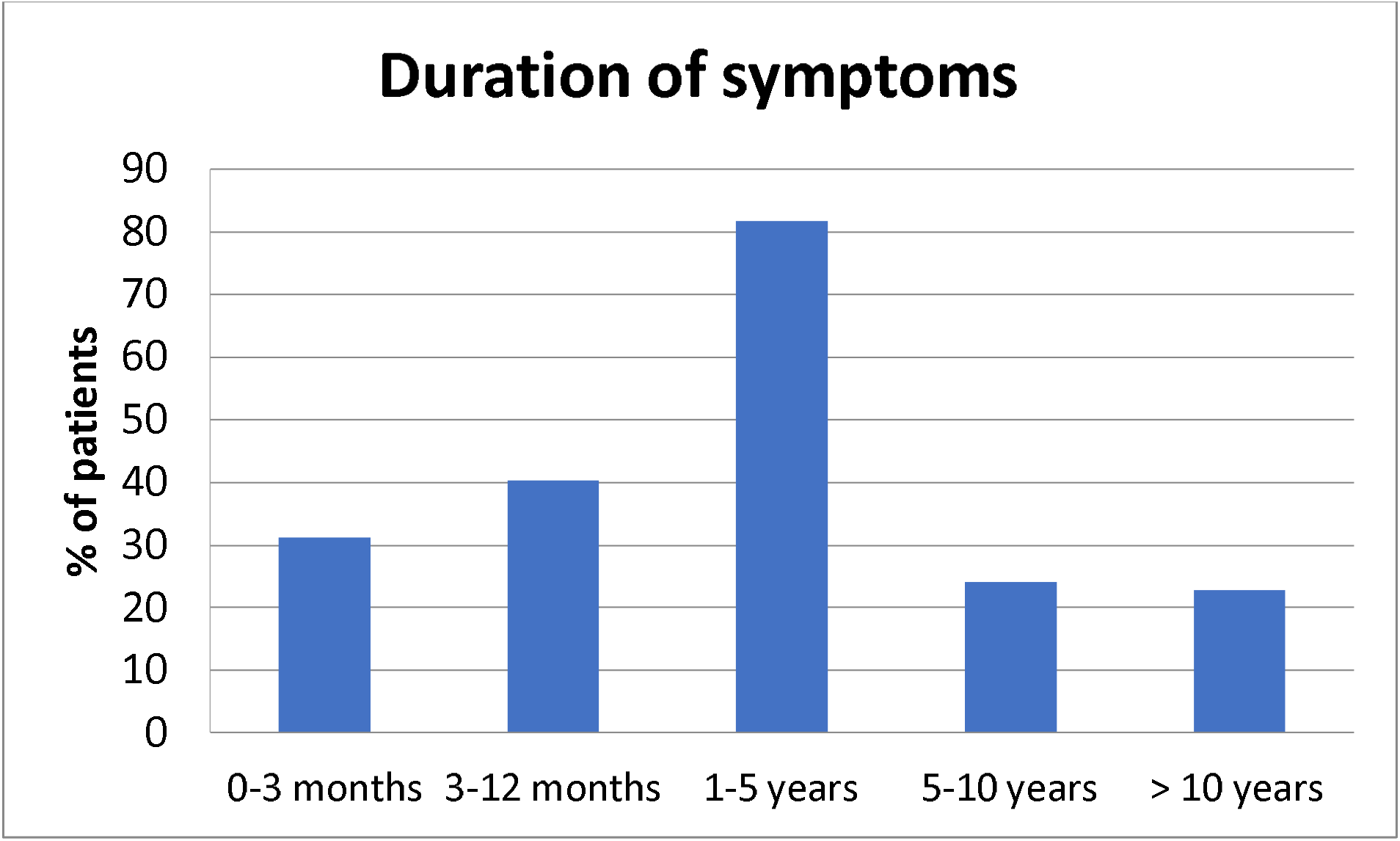
Symptoms duration in the whole group of patients.

### Comparisons between groups (see Table 2 and supplementary table 1)

#### Comparisons between functional motor weakness patients and functional movement disorders patients

Patients with functional motor weakness were significantly younger at TMS treatment than patients with functional movement disorders (t=-3.75, p<0.001). However, disease duration before TMS treatment was shorter in patients with functional motor weakness (t=-6.01, p<0.001), and there was no significant difference between both groups regarding age at symptoms onset (t=-1.077, p=0.27). There was no significant difference between the two groups in terms of education level, occupation, medical somatic history, psychiatric comorbidities, self-reported trauma history or symptoms onset (see table 2).

**Table 2.** Demographic and clinical characteristics of patients according to the functional motor disorder phenotype.

|  |  | Functional Weakness | Functional Movement Disorder | Statistics |
| --- | --- | --- | --- | --- |
| N |  | 241 | 241 |  |
| Age at TMS in years, Mean (SD) |  | 37.2 (13.7) | 42.2 (16.1) | p<0.001 |
| Gender (Women in % (N)) |  | 76.3% (184) | 70.9% (171) | ns |
| Education (≥ in % (N)) |  | 42.3% (102) | 48.9% (113) | ns |
| occupation | Active | 25.7% (62) | 20.3% (49) | ns |
|  | medical reason | 57.7% (139) | 58.9% (142) |  |
|  | non-medical reasons | 16.6% (40) | 20.7% (50) |  |
| Age at onset in years (Mean (SD) / Median) |  | 34.6 (13.0) / 36 | 36.1 (16.5) / 35 | ns |
| Duration in months (Mean (SD)/Median) |  | 30.1 (43.1) / 15 | 73.8 (104.1) / 37 | p<0.001 |
| Onset | Acute | 50.2% (121) | 44.4% (107) | ns |
|  | Subacute | 44.8% (108) | 47.3% (114) |  |
|  | Chronic | 5.0% (12) | 8.3% (20) |  |

#### Comparisons between FMDs’ phenotypes

There was no significant difference in term of age at TMS, age at symptoms onset, disease duration, sex ratio, educational level, occupation, medical somatic history, psychiatric comorbidities, self-reported trauma history or symptoms installation between the 5 phenotypes of FMD. Nonetheless, patients with parkinsonism tended to be older at disease onset (F=1.47; p=0.21). Likewise, patients with dystonia and tremor tended to have larger symptoms durations than patients with parkinsonism, jerky dystonia and stereotypies (F=1.43; p= 0.22).

#### Topography of symptoms

Location of symptoms is detailed in Table 3 and supplementary table 3. In both FMD and motor deficit, bilateral symptoms were predominant. However, symptoms were more likely to be bilateral in FMD (FMD: n=133/241 (55.2%); weakness: n= 116/241 (48.1%)), and more likely to be left-sided in weakness, (left sided in FMD: n=53/241 (22%); weakness: n=83/241 (34.4%)) (χ^2^=9.52; p= 0.009). Additionally, Statistics (χ^2^=125.7; p< 0.001) revealed a significantly higher proportion of left-sided symptoms for the lower limbs and a right-sided predominance of symptoms for upper limbs amongst patients with functional motor weakness and dystonia (with dystonia being more often monomelic regarding the lower limbs and targeting especially the head and neck compared to the motor weakness group). Tremor, parkinsonism and jerky dystonia symptoms were mainly bilateral. Finally, lower limbs symptoms were predominant only in patients with functional motor weakness (χ^2^=89.50; p< 0.001).

**Table 3.** Topography of symptoms according to the functional motor disorder phenotype.

|  | FMD<br>(n(%)) | Motor deficit<br>(n(%)) | Statistics |
| --- | --- | --- | --- |
| Bilateral (N(%)) | 133 (55.2) | 116 (48,1) | $\chi^2=9.52$; p= 0.009 |
| Right (N(%)) | 55 (22.8) | 42 (17,4) |  |
| Left (N(%)) | 53 (22) | 83 (34,4) |  |
| **Total (N(%))** | **241 (100)** | **241 (100)** |  |

## Discussion

### Demographic characteristics

We present here demographic and clinical data of a large cohort of patients with FMDs. The majority of patients were women with a median age at the symptoms onset of 35 years old, with a wide age range. These demographic characteristics are similar to those of previous smaller cohorts of adults suffering from FMDs, which reported a proportion of women between 48 and 84% and a mean age at onset between 29.7 and 46.9 years old (Factor, Podskalny, et Molho 1995; Binzer, Andersen, et Kullgren 1997; Schrag et al. 2004; Jon Stone, Warlow, et Sharpe 2010; Pandey et Koul 2017; Baizabal-Carvallo et Jankovic 2020; Tinazzi et al. 2020; Anderson et al. 2007; Gelauff et al. 2020)(see table 4). It should be noted that extreme ages were under-represented, since wee did not include children, and older patients may be under diagnosed. Patients had relatively long disease duration, with a mean duration of 52 months in our cohort that may be explained by a recruitment bias because the Pitié Salpêtrière neurology department is a tertiary center, notably regarding movement disorders.

**Table 4.** Previous reported cohorts of patients suffering from FMD. NA: not available

| reference | number of cases | phenotypes | Acute Onset | mean age | mean age at onset | sex ratio (%F) | mean duration (months) | % not working/ illness* |
| --- | --- | --- | --- | --- | --- | --- | --- | --- |
| Factor et al. 95 | 28 | Movement disorders | 54% | 49.7 | 46.9 | 60.7% | 33 | NA |
| Binzer et al 1997 | 30 | weakness | all patients <3 months | 38.8 | NA | 60% | <3 | NA |
| Schrag et al 2004 | 103 | fixed dystonia | NA | 33 | 29.7 | 84% | 39.6 | NA |
| Anderson et al 2007 | 66 | movement disorders | NA | 49.6 | 46.5 | 65.2% | 37.2 | 47%/NA |
| Stone et al 2010 | 107 | weakness | 61% | 39 | 38.1 | 79% | 11 | 68%/65% |
| Pandey et al 2017 | 33 | movement disorders | 75.75% | 37.5 | 36.7 | 48% | 10.2 | NA |
| Baizabal-Carvallo et al 2019 | 196 | movement disorders | NA | 39.6 | 36 | 70% | 43.2 | NA |
| Tinazzi et al 2020 | 410 | motor symptoms | 70.7% | 47 | 41,5 | 71% | 66 | NA |
| gelauff et al 2020 | 160 | motor symptoms | 68% in < 6h or at wakening | 48 | 46 | 71% | 24 | 74%/56% |
| Our cohort | 482 | Motor symptoms | 90% | 39.7 | 35.4 | 73.7% | 52 | 77%/58% |

\* illness: not working because of the FMD.

### Onset and clinical characteristics

Amongst movement disorders, tremor and dystonia were the most prevalent presentations, this has also been described in previous cohorts of movement disorders (Factor, Podskalny, et Molho 1995; Pandey et Koul 2017; Baizabal-Carvallo et Jankovic 2020; Tinazzi et al. 2020; Anderson et al. 2007)

We found no significant demographic differences between functional weakness and functional movement disorders. Another recent study failed to find differences between groups of functional motor symptoms, regarding demo-graphics and mode of onset. The authors concluded that the large overlap in symptoms contributes to the hypothesis of shared underlying mechanisms of functional motor disorders. Alternatively, the absence of significant difference might due to a lack of power in this study, and multicenter cohorts, as well as meta-analyses, would be necessary to better characterize the differences and commonalities of patients of to different FMD phenotypes. To note, there was a tendency towards older age at onset in movement disorders, notably in patients with functional parkinsonism (mean: 48.3 years old). In a recent literature review, the mean age at onset in functional parkinsonism was 45.6 years (Ambar Akkaoui et al. 2020). On the contrary, age at onset was lower in functional dystonia (mean: 35.2), and this is also in agreement with a previous study that found age at onset of 29.6 years old in functional dystonia. It is interesting to note this age difference is similar to what is observed in so-called “organic” diseases, where dystonia affects younger patients than Parkinson’s disease. This raises the question of similar pathophysiology in functional and organic disorders.

In our cohort, weakness affected more the left side, notably when the lower limbs were affected, whereas movement disorders were more likely to be bilateral. The lateralization of FMD has been a subject of debate (J. Stone et al. 2002), although this was not the aim of the study, our results raise the question of lateralization of functional weakness. Further studies would be necessary to confirm these results and to determine the physiopathological explanation to this lateralization.

An acute or subacute onset was reported in 95 % of patients in our cohort. Acute onset is often reported in FMD, and was noticed in the majority of patients in previous cohorts of FMD (Jon Stone, Warlow, et Sharpe 2012; Tinazzi et al. 2020; Factor, Podskalny, et Molho 1995; Pandey et Koul 2017). It is usually considered as a clue for the diagnosis of functional movement disorders (Espay et al. 2018). In a previous study, panic and dissociative symptoms were associated with a sudden onset of FMD, however, the reasons why symptoms occur suddenly are still not clearly elucidated.

### Comorbidities

In our cohort, 18.5 % had a neurological comorbidity, and 66.4% had a psychiatric comorbidity. These figures were comparable to those described in an Italian cohort of 410 patients with FMD, where 17% had a comorbidity of neurological disorder, and 40% had a psychiatric comorbidity. To note, a significant proportion of patients had no psychiatric comorbidity or traumatic events history. This is in agreement with the exclusion of psychological factors in the latest diagnostic criteria of FND (Jon Stone et al. 2011, 5).

### Disability

In our cohort, 77% of patients were not working and 58% were not working because of FND. This data is not available in most other cohorts. Unemployment was reported in two previous cohorts of FMD patients: unemployment rate was 47% in a cohort of 66 functional movement disorders patients regardless of the reason of unemployment (Anderson et al. 2007), and in a cohort of 107 patients with functional weakness, 65% of the patients were not working because of their symptoms (Jon Stone, Warlow, et Sharpe 2010). These proportions are higher than in a previous study of neurology outpatients with symptoms ‘unexplained by organic disease’, where 297 out of 1144 (26%) patients were not working because of their symptoms (Carson et al. 2011; Jon Stone, Warlow, et Sharpe 2010). More generally, it has been shown that patients with medically unexplained symptoms or somatoform disorders have a higher risk of sick leave and disability pension award during follow up (Rask et al. 2015).

Our results underline the socio-economical costs of FMD and stress out the urge for a better diagnostic procedure and management.

## Data Availability

All data can be provided on request to the corresponding author

